# Development and Validation of Prediction Models for Sentinel Lymph Node Status Indicating Postmastectomy Radiotherapy in Breast Cancer: a Population-Based Study of 18 185 Women

**DOI:** 10.1101/2024.02.21.24303165

**Authors:** Miriam Svensson, Pär-Ola Bendahl, Sara Alkner, Emma Hansson, Lisa Rydén, Looket Dihge

**Affiliations:** Department of Clinical Sciences, Division of Surgery, Lund University, Lund, Sweden; Department of Clinical Sciences, Division of Oncology and Pathology, Lund University, Lund, Sweden; Department of Hematology, Oncology and Radiation Physics, Skåne University Hospital, Lund, Sweden; Department of Plastic Surgery, Institute of Clinical Sciences, Sahlgrenska Academy, University of Gothenburg, Gothenburg, Sweden; Region Västra Götaland, Sahlgrenska University Hospital, Department of Plastic Surgery, Gothenborg Sweden; Department of Surgery, Skåne University Hospital, Malmö, Sweden; Department of Plastic and Reconstructive Surgery, Skåne University Hospital, Malmö, Sweden

**Keywords:** Breast Neoplasms, Sentinel Lymph Node, Radiotherapy, Breast Reconstruction, Nomograms, Clinical decision support

## Abstract

**Background:** Postmastectomy radiotherapy (PMRT) impairs the outcome of immediate breast reconstruction (IBR) in patients with breast cancer, and the sentinel lymph node (SLN) status is crucial in evaluating the need for PMRT. This study aimed to develop models to preoperatively predict the risk for SLN metastasis indicating the need for PMRT.

**Methods:** Women diagnosed with clinically node-negative (cN0) T1-T2 breast cancer from January 2014 to December 2017 were identified within the Swedish National Quality Register for Breast Cancer. Nomograms for nodal prediction based on preoperatively accessible patient and tumor characteristics were developed using adaptive LASSO logistic regression. The prediction of ≥1 and >2 SLN macrometastases (macro-SLNMs) adheres to the current guidelines on use of PMRT and reflects the exclusion criteria in ongoing clinical trials aiming to de-escalate locoregional radiotherapy in patients with 1-2 macro-SLNMs, respectively. Predictive performance was evaluated using area under the receiver operating characteristic curve (AUC) and calibration plots.

**Results:** Overall, 18 185 women were grouped into training (*n* =13 656) and validation (*n* = 4529) cohorts. The well-calibrated nomograms predicting ≥1 and >2 macro-SLNMs displayed AUCs of 0.708 and 0.740, respectively, upon validation. By using the nomogram for ≥1 macro-SLNMs, the risk could be updated from the pre-test population prevalence 13% to the post-test range 2%-75%.

**Conclusion:** Nomograms based on routine patient and tumor characteristics could be used for prediction of SLN status that would indicate PMRT need and assist the decision-making on IBR for patients with cN0 breast cancer.

## Introduction

Breast reconstructive surgery improves the quality of life of patients with breast cancer undergoing mastectomy^1, 2^. According to international guidelines, all patients undergoing mastectomy should be counseled about reconstructive options. To evaluate the optimal timing and type of breast reconstruction, risk factors for postoperative complications must be considered, such as smoking^3^, obesity^4^, and diabetes^5^. Specifically, postmastectomy radiotherapy (PMRT) is recognized to impact outcomes in those receiving immediate breast reconstruction (IBR)^6–10^. Common complications involve capsular contracture and loss of implant following immediate implant-based reconstructions^6, 7^, while tissue necrosis is a common complication associated with immediate autologous reconstructions and PMRT^11^. Therefore, evaluation of the need for PMRT is essential to support patients with breast cancer and health-care providers in making informed decisions on breast reconstructive surgery and IBR in particular.

The decision on PMRT is based on axillary lymph node metastasis, tumor size of >50 mm or involved resection margins^12–14^. Sentinel lymph node biopsy (SLNB) is the gold standard for axillary nodal staging in patients with clinically node-negative (cN0) invasive breast cancer, and all patients with ≥1 SLN macrometastases (macro-SLNMs) (>2.0 mm) receive PMRT, according to current guidelines^12–14^. Ongoing randomized clinical trials are examining the possibility of omitting locoregional radiotherapy in patients with 1–2 macro-SLNMs^15–17^. However, for those with a heavy nodal disease burden, PMRT remains advisable. Consequently, the identification of patients with >2 macro-SLNMs is crucial. A non-invasive tool for predicting SLN status could be beneficial in preoperatively identifying patients at risk of postoperative complications associated with the requirement for PMRT following IBR. Although there are several nomograms for predicting axillary node status^18–20^, most of them include variables that are not routinely available in a preoperative setting. Moreover, predictive tools to identify patients with cN0 breast cancer at high risk of macro-SLNM indicating the need for PMRT, are still lacking.

This study aims to develop and validate nomograms for risk stratification of SLN metastasis indicating the need for PMRT in patients with cN0 breast cancer using only routine clinical patient and tumor characteristics. The primary endpoint is the prediction of ≥1 macro-SLNMs, according to current guidelines on the use of PMRT. The secondary endpoint is the prediction of >2 macro-SLNMs that adhere to the exclusion criteria of ongoing randomized controlled trials aiming to de-escalate the use of locoregional radiotherapy in patients with low nodal metastatic burden. Adaptive LASSO logistic regression is used to minimize the risk of overfitting the models. To our knowledge, this is the first study to present nomograms that stratify the risk for SLNM, indicating the need for PMRT in patients with cN0 breast cancer.

## Methods

### Study population

This retrospective study was conducted using data from the Swedish National Quality Register for Breast Cancer (NKBC), a prospectively maintained, population-based register with almost 100% completeness when cross-linked to the Swedish Cancer Register^21^. All women diagnosed with invasive breast cancer in Sweden from January 2014 to December 2017, primarily treated with surgery, were identified. The exclusion criteria were: bilateral breast cancer, neoadjuvant chemotherapy, ductal carcinoma *in situ* (DCIS), tumor size >50 mm or unknown tumor size, stage IV breast cancer, palpable axillary lymphadenopathy, absent or incongruent data on axillary nodal status and omission of SLNB. The dataset was split into a training cohort and a validation cohort. The training cohort comprised patients diagnosed in 2014-2016, while the remaining patients constituted the temporal validation cohort.

For all included patients, SLNB was the primary axillary staging procedure for evaluation of axillary nodal status, and nodal metastases were categorized into macrometastases if a metastatic deposit >2.0 mm was displayed^22^. For the prediction of >2 macro-SLNMs, patients with macrometastases in at least two sentinel nodes and with any additional lymph node metastasis identified by SLNB or by completion axillary lymph node dissection, were included.

This research was performed in accordance with the Declaration of Helsinki and followed the STROBE and TRIPOD guidelines for reporting observational studies^23, 24^. The study was approved by the Swedish Ethical Review Authority (2019–02139). Written informed consent for participation was not required for this register-based study in accordance with the national legislation and institutional requirements. The study was registered in the ISRCTN database (ISRCTN 14341750).

### Clinicopathological predictors

Based on previous studies on variables associated with SLN status^25, 26^, the following candidate predictors were evaluated: age at diagnosis, tumor size, Nottingham histological grade, histological type, estrogen receptor (ER) status, progesterone receptor (PR) status, amplification of human epidermal growth factor receptor 2 (HER2), and multifocality. Patient age has been reported to have a non-linear association with nodal status, with the lowest prevalence of lymphatic involvement for those around 70 years^27^. Therefore, patients were categorized into the following age groups: ≤65, 66–75 and >75 years. Multifocality was defined as the presence of ≥2 foci of invasive breast cancer within the same breast separated by benign tissue on the histological examination of the excised section^28^, and tumor size was defined as the greatest dimension of the largest invasive focus. For ER and PR status, ≥1% stained nuclei by immunohistochemistry (IHC) were considered positive according to the definitions of the European Society for Medical Oncology (ESMO)^12^. To evaluate HER2 status, IHC and *in situ* hybridization (ISH) were performed, and tumors with IHC 3+ scoring and/or positive ISH-test were regarded as HER2-positive. The histological type was categorized into three groups: invasive carcinoma of no special type (NST), invasive lobular carcinoma (ILC), and other invasive carcinoma. Mixed types were excluded from the analyzes.

### Statistical analysis

Univariable logistic regression analysis was used to explore the unadjusted associations between each candidate predictor and the two endpoints in the training cohort. Adaptive LASSO logistic regression^29^ was then applied to select the most important predictors. LASSO regression is a commonly used machine learning technique for variable selection and model development, minimizing the risk of overfitting by forcing the absolute sizes of the regression coefficients of the standardized predictors to be bounded by a penalty factor, λ. The adaptive form of LASSO leads to even more parsimonious models than the standard LASSO^30,31^. To determine the optimal value of λ, we used 10-fold cross-validation in the training cohort. Cases with missing values for ≥1 candidate predictors were removed from the analyses. Two nomograms for predicting ≥1 and >2 macro-SLNMs, respectively, were developed based on the results from the adaptive LASSO regression analyses.

To evaluate each nomogram’s discriminatory ability, the area under the receiver operating characteristic (ROC) curve (AUC) was calculated in the development and validation cohorts. Model accuracy was assessed in the validation cohort using calibration plots (graphical) and calibration slope and intercept (numerical). For comparison purposes, corresponding prediction models based on backward stepwise regression with uniform bootstrap-based shrinkage^32^ were developed and are provided in the supplementary section (Supplemental Methods). All analyses were performed using STATA (StataCorp. 2021. *Stata Statistical Software: Release 17.* College Station, TX: StataCorp LLC).

## Results

### Patient and tumor characteristics

From January 2014 to December 2017, 23 256 breast cancer tumors, primarily treated with surgery, were registered in NKBC (Fig. S1). The final study cohort consisted of 18 185 patients with breast cancer who met the eligibility criteria. Of these, 13 656 were diagnosed between 2014 and 2016, constituting the training cohort, and 4529 were diagnosed in 2017, constituting the temporal validation cohort. Patient and tumor characteristics of the training and validation cohorts were comparable (Table 1). Overall, 2409 patients (13%) displayed ≥1 macro-SLNMs, and 278 (2%) displayed >2 macro-SLNMs. The overall median age at diagnosis was 64 years, and the median tumor size was 15 mm. Most patients displayed unifocal, hormone receptor-positive, HER2-negative, grade II carcinoma of NST.

**Table 1.**
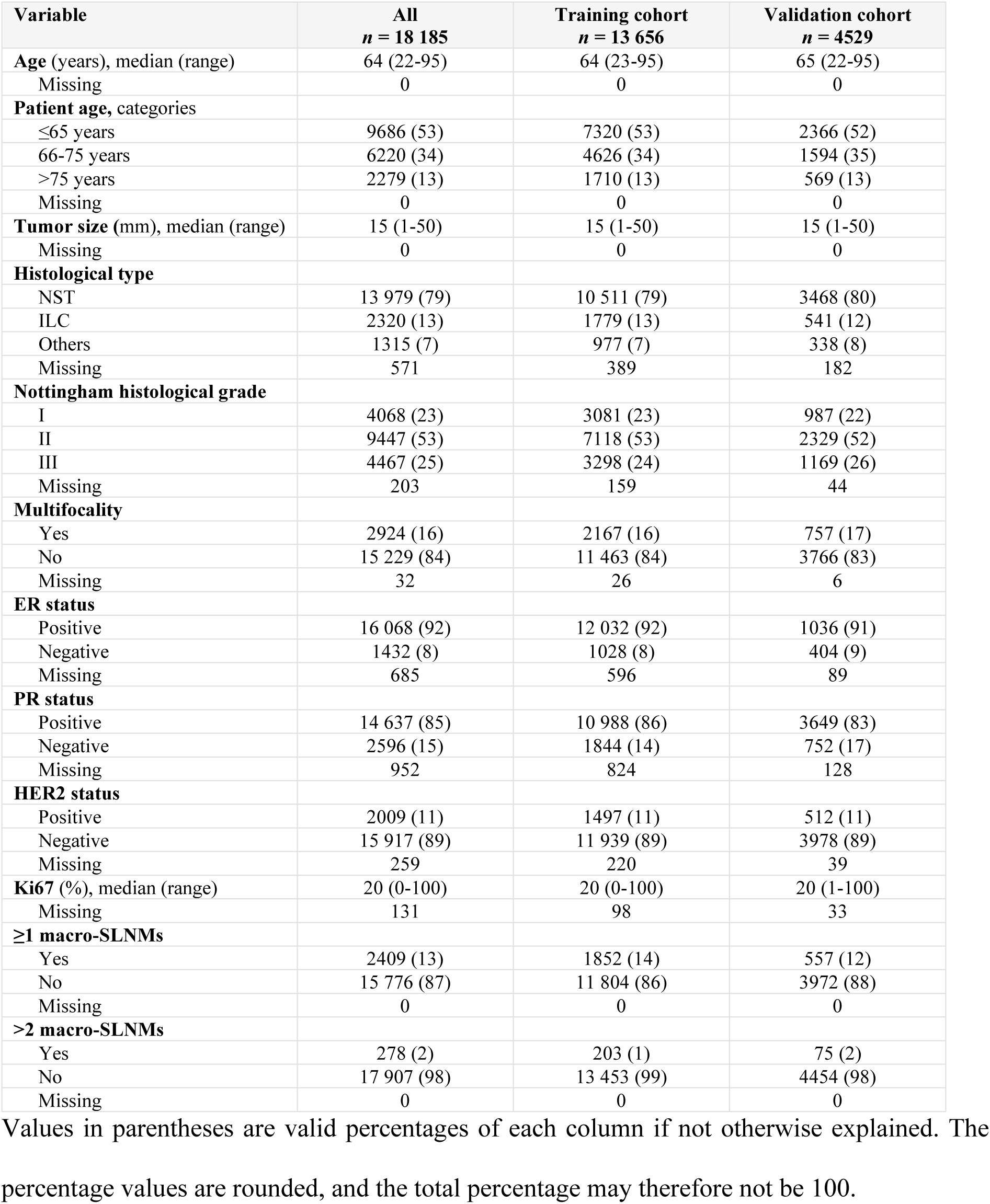
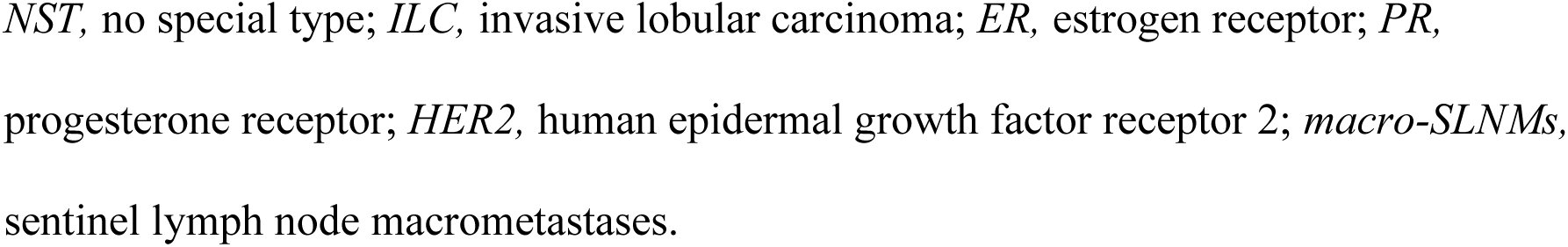
Patient and tumor characteristics of the training and validation cohorts.

| Variable | All<br>*n* = 18 185 | Training cohort<br>*n* = 13 656 | Validation cohort<br>*n* = 4529 |
| --- | --- | --- | --- |
| **Age** (years), median (range) | 64 (22-95) | 64 (23-95) | 65 (22-95) |
| Missing | 0 | 0 | 0 |
| **Patient age**, categories |  |  |  |
| ≤65 years | 9686 (53) | 7320 (53) | 2366 (52) |
| 66-75 years | 6220 (34) | 4626 (34) | 1594 (35) |
| >75 years | 2279 (13) | 1710 (13) | 569 (13) |
| Missing | 0 | 0 | 0 |
| **Tumor size** (mm), median (range) | 15 (1-50) | 15 (1-50) | 15 (1-50) |
| Missing | 0 | 0 | 0 |
| **Histological type** |  |  |  |
| NST | 13 979 (79) | 10 511 (79) | 3468 (80) |
| ILC | 2320 (13) | 1779 (13) | 541 (12) |
| Others | 1315 (7) | 977 (7) | 338 (8) |
| Missing | 571 | 389 | 182 |
| **Nottingham histological grade** |  |  |  |
| I | 4068 (23) | 3081 (23) | 987 (22) |
| II | 9447 (53) | 7118 (53) | 2329 (52) |
| III | 4467 (25) | 3298 (24) | 1169 (26) |
| Missing | 203 | 159 | 44 |
| **Multifocality** |  |  |  |
| Yes | 2924 (16) | 2167 (16) | 757 (17) |
| No | 15 229 (84) | 11 463 (84) | 3766 (83) |
| Missing | 32 | 26 | 6 |
| **ER status** |  |  |  |
| Positive | 16 068 (92) | 12 032 (92) | 1036 (91) |
| Negative | 1432 (8) | 1028 (8) | 404 (9) |
| Missing | 685 | 596 | 89 |
| **PR status** |  |  |  |
| Positive | 14 637 (85) | 10 988 (86) | 3649 (83) |
| Negative | 2596 (15) | 1844 (14) | 752 (17) |
| Missing | 952 | 824 | 128 |
| **HER2 status** |  |  |  |
| Positive | 2009 (11) | 1497 (11) | 512 (11) |
| Negative | 15 917 (89) | 11 939 (89) | 3978 (89) |
| Missing | 259 | 220 | 39 |
| **Ki67** (%), median (range) | 20 (0-100) | 20 (0-100) | 20 (1-100) |
| Missing | 131 | 98 | 33 |
| **≥1 macro-SLNs** |  |  |  |
| Yes | 2409 (13) | 1852 (14) | 557 (12) |
| No | 15 776 (87) | 11 804 (86) | 3972 (88) |
| Missing | 0 | 0 | 0 |
| **>2 macro-SLNs** |  |  |  |
| Yes | 278 (2) | 203 (1) | 75 (2) |
| No | 17 907 (98) | 13 453 (99) | 4454 (98) |
| Missing | 0 | 0 | 0 |

Values in parentheses are valid percentages of each column if not otherwise explained. The percentage values are rounded, and the total percentage may therefore not be 100.
*NST*, no special type; *ILC*, invasive lobular carcinoma; *ER*, estrogen receptor; *PR*, progesterone receptor; *HER2*, human epidermal growth factor receptor 2; *macro-SLNs*, sentinel lymph node macrometastases.

### Variable selection and prediction model development using adaptive logistic LASSO regression

The results from the univariable logistic regression analyses are presented in Table S1. Only patients with complete information on all candidate predictors were included in the adaptive LASSO logistic regression analyses (*n* = 12 168, 89%). Patient and tumor characteristics of those included in the analyses and those removed due to any missing value of the candidate predictors are presented in Table S2. For prediction of ≥1 macro-SLNMs, the adaptive LASSO regression-based prediction model identified patient age, tumor size, multifocality, histological type, histological grade, ER, PR, and HER2 status as predictors. Tumor size emerged as the most important predictor, followed by multifocality (Fig. S2a and S2b).

For the prediction of >2 macro-SLNMs, adaptive LASSO regression identified the following predictors: tumor size, histological grade, multifocality, patient age (>65 years *vs.* ≤65 years), and histological type (ILC *vs.* NST or others) (Fig. S2c and S2d). Tumor size was also the strongest predictor in this model.

The regression coefficients from the two adaptive LASSO regression analyses are presented in Table 2. Penalization of the coefficients was applied to minimize the risk of overfitting. Along with the estimated intercept, these coefficients constitute the function used for outcome prediction. A positive coefficient indicates that the variable increases the predicted probability, and a negative coefficient indicates that the variable decreases the predicted probability. The retained predictors and their corresponding regression coefficients in the two supplementary prediction models based on backward stepwise regression with bootstrap uniform shrinkage are presented in Supplemental Results and Table S3.

**Table 2.** Penalized regression coefficients for prediction of ≥1 and >2 sentinel lymph node macrometastases (macro-SLNMs) identified by adaptive LASSO regression (*n* = 12 168)

| Variable | Prediction of $\geq 1$ macro-SLNMs<br>Penalized coefficients | Prediction of $> 2$ macro-SLNMs<br>Penalized coefficients |
| --- | --- | --- |
| **Age (years)** |  |  |
| $\leq 65$ | 0 (reference) | |
| 66-75 | -0.276 |  |
| $> 75$ | -0.128 | |
| **Age (years) ($> 65$ vs. $\leq 65$)** |  | -0.171 |
| **Tumor size (mm)** | 0.068 | 0.076 |
| **Histological type** |  |  |
| NST | 0 (reference) |  |
| ILC | -0.270 |  |
| Others | -0.658 |  |
| **Histological type (ILC vs. NST or others)** |  | 0.070 |
| **Histological grade** |  |  |
| I | 0 (reference) | 0 (reference) |
| II | 0.359 | 0.743 |
| III | 0.489 | 0.813 |
| **Multifocality (multifocal vs. unifocal)** | 0.643 | 0.489 |
| **ER status (pos vs. neg)** | 0.225 |  |
| **PR status (pos vs. neg)** | 0.122 |  |
| **HER2 status (pos vs. neg)** | -0.078 |  |
| **Constant** | -3.669 | -6.451 |

Binary coding was used for categorical variables with two levels and two binary so-called dummy variables for categorical variables with three levels.
*NST*, ductal carcinoma of no special type; *ILC*, invasive lobular carcinoma; *ER*, estrogen receptor; *PR*, progesterone receptor; *HER2*, human epidermal growth factor receptor 2.

### Nomogram development

The results from the adaptive LASSO logistic regression analyses were used to develop Nomograms I and II, predicting ≥1 and >2 macro-SLNMs, respectively (Fig. 1). The penalized regression coefficients were transformed into specific scores on scales ranging from 0 to 10. By summarizing all scores in one nomogram, the patient’s total score can be applied to a separate scale to predict the presence of ≥1 macro-SLNMs and >2 macro-SLNMs, respectively.

**Figure 1.**
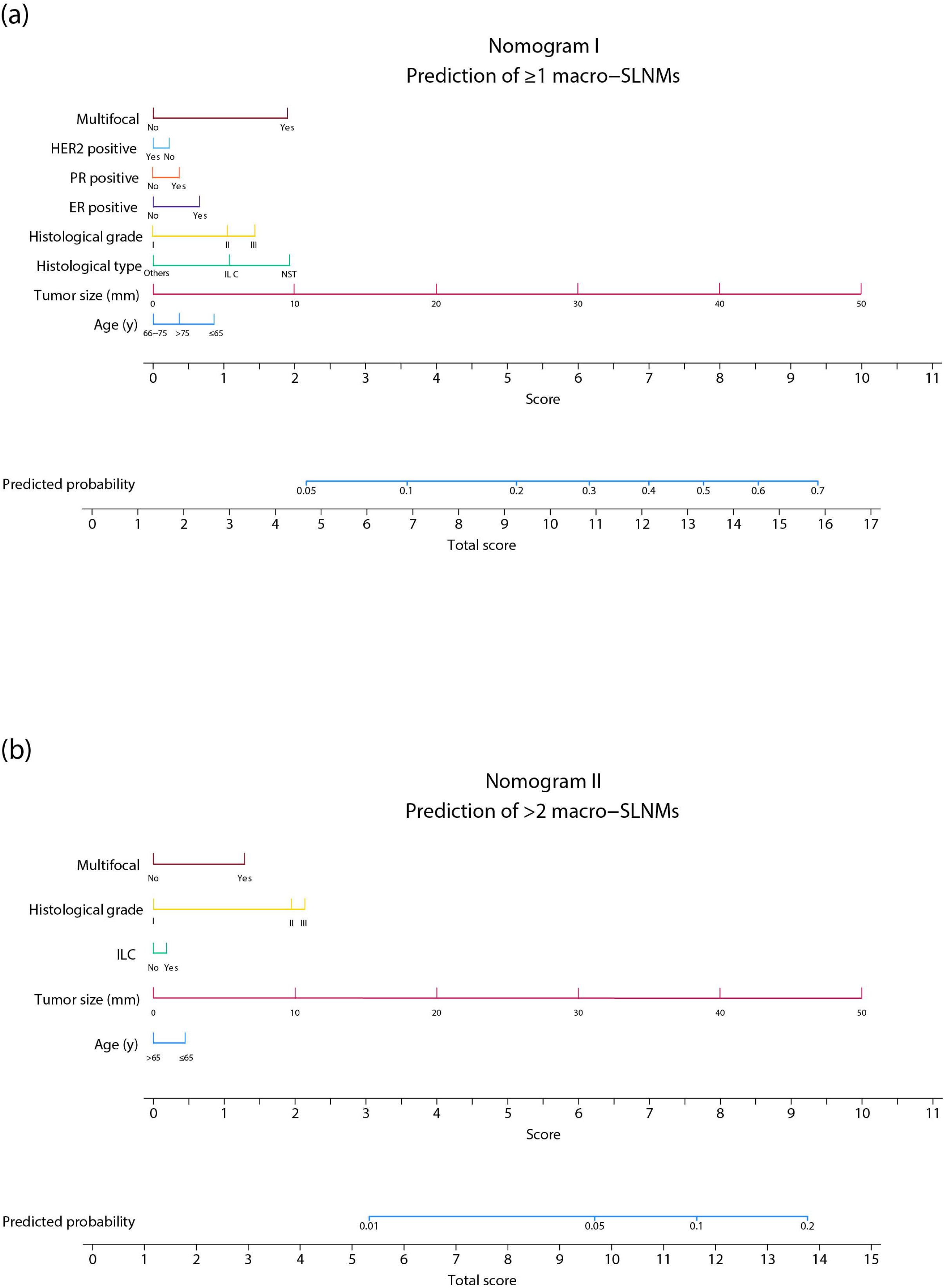
Nomograms predicting sentinel lymph node (SLN) status indicating the need for postmastectomy radiotherapy (PMRT): (**a**) probability of ≥1 SLN macrometastases (macro-SLNMs) according to current guidelines on use of PMRT; and (**b**) probability of >2 macro-SLNMs according to endpoints of ongoing clinical trials aiming to de-escalate the use of irradiation. *HER2,* human epidermal growth factor receptor 2; *PR,* progesterone receptor; *ER,* estrogen receptor; *ILC,* invasive lobular carcinoma; *NST,* ductal carcinoma of no special type.

### Prediction model performance

The ROC curves and calibration plots illustrating the discriminatory ability and the accuracy of the two prediction models in the validation cohort are presented in Fig. 2. For Nomogram I, the AUC value was 0.720 (95% confidence interval CI, 0.707–0.733) in the training cohort and 0.708 (0.684–0.731) in the validation cohort, respectively. As illustrated by the calibration plot, the nomogram displayed good agreement between the predicted and observed prevalence of macro-SLNM in the validation cohort. The calibration slope and intercept were estimated to be 1.030 and −0.108, respectively. These estimates are close to the optimal values, which are 1.000 and 0.000, respectively. When applying the nomogram to the overall study cohort, the individually predicted probability of ≥1 macro-SLNMs ranged from 2% for some low-risk patients up to 75% for some high-risk patients (Fig. 3).

**Figure 2.**
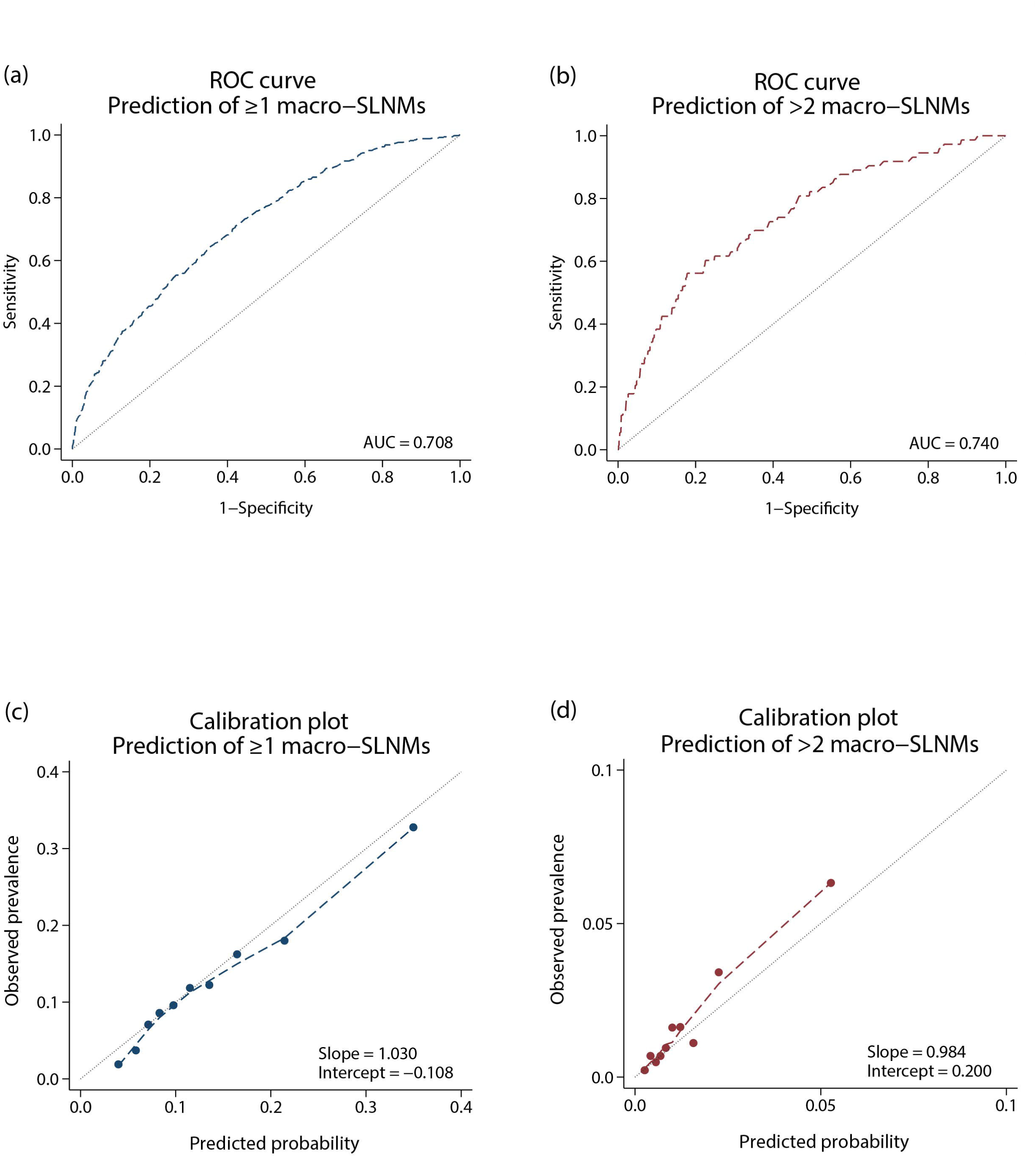
Performance of the nomograms in the temporal validation cohort. Receiver operating characteristic (ROC) curves representing the discriminatory ability for (**a**) Nomogram I, predicting ≥1 sentinel lymph node macrometastases (macro-SLNMs), and (**b**) Nomogram II, predicting >2 macro-SLNMs. The Calibration plots illustrating the agreement between the observed prevalence and the predicted probability of (**c**) ≥1 macro-SLNMs and (**d**) >2 macro-SLNMs, respectively, show good calibration, i.e., the predictions are not systematically biased. *AUC,* area under the curve.

**Figure 3.**
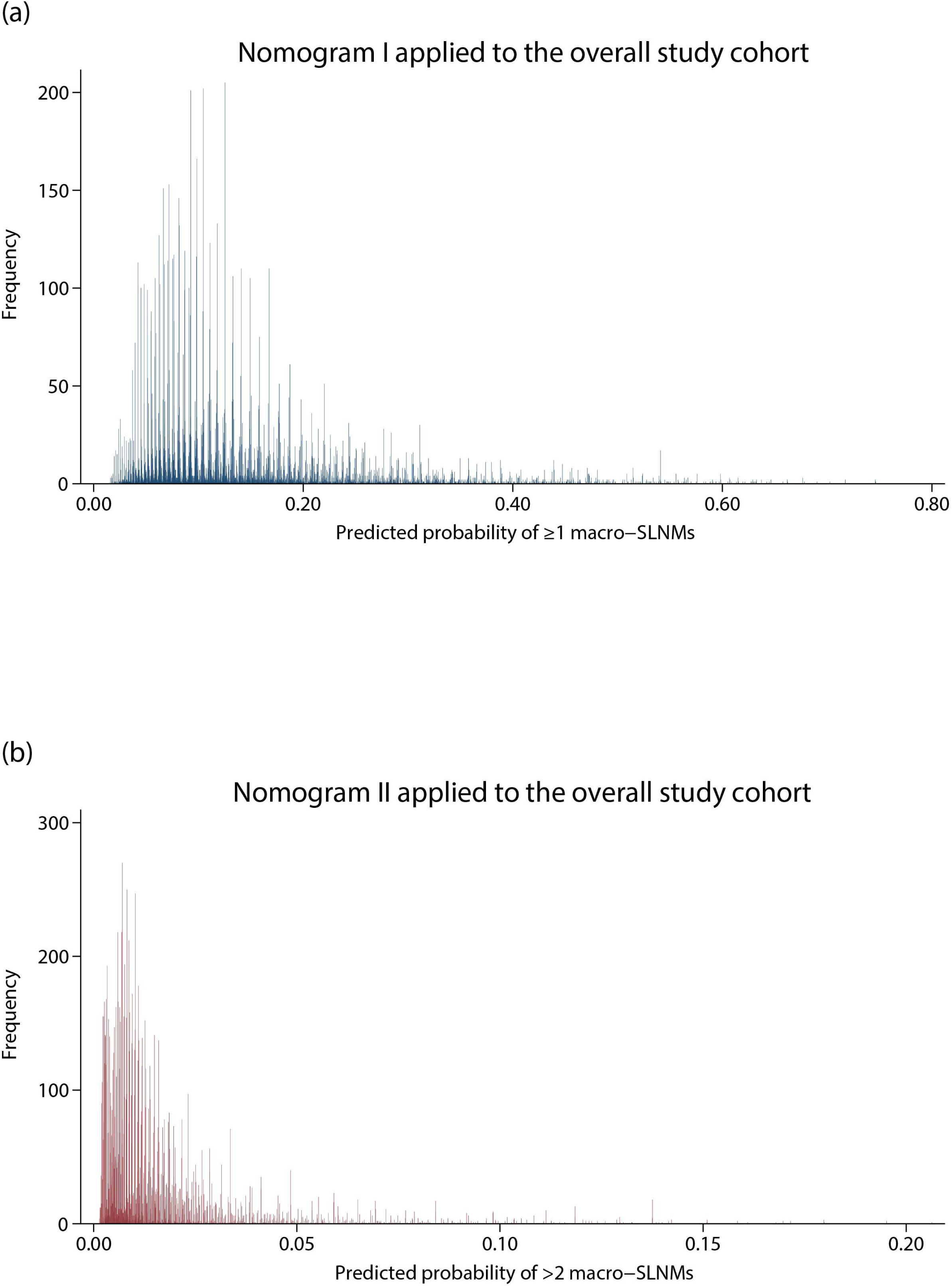
Individual predictions of ≥1 and >2 sentinel lymph node macrometastases (macro-SLNMs) when applying the nomograms to the overall study cohort. (**a**) For Nomogram I, the predicted probability of ≥1 macro-SLNMs ranged from 2% for some low-risk patients up to 75% for some high-risk patients. (**b**) Likewise, the predicted probability of >2 macro-SLNMs ranged from <1% to 21% when applying Nomogram II to each patient in the overall study cohort.

For Nomogram II, the AUC values in the training and validation cohorts were 0.775 (0.743–0.807) and 0.740 (0.682–0.799), respectively (Fig. 2b). Similarly, this nomogram was well-calibrated with close approximation between the observed prevalence and predicted probability in the validation cohort (Fig. 2d), with a calibration slope and intercept of 0.984 and 0.200, respectively. The individually predicted probability of >2 macro-SLNMs ranged from <1% to 21% in the overall study cohort, with a mean of 2%. AUC values and model calibration for the two supplementary models based on logistic regression are presented in Supplemental results and Fig. S3.

## Discussion

This study presents nomograms based on routine clinical patient and tumor characteristics for preoperative risk stratification of macro-SLNM indicating the need for PMRT. The chosen endpoints are according to the current guidelines on the use of PMRT and reflect ongoing clinical trials aiming to de-escalate locoregional irradiation in patients with low nodal metastatic burden. The nomograms displayed good discriminative ability, were well-calibrated, and could be used to increase or decrease the preoperative likelihood of macro-SLNM. For some high-risk patients, the estimated risk of ≥1 macro-SLNMs increased from the study population prevalence of 13% to 75% when using Nomogram I, while for others, the estimated risk decreased to 2%. This way, the nomograms can assist patients with breast cancer and health-care providers in making informed decisions regarding reconstructive surgery.

To evaluate nodal status indicating the need for PMRT before a decision on IBR, intraoperative SLN staging was previously performed using imprint cytology or frozen sections^33^. However, besides being time-consuming, intraoperative evaluation of the SLN status inevitably leaves the patient with some degree of preoperative uncertainty regarding the nodal status and suitability of IBR. Additionally, it complicates surgical planning, impacting the required time in the operating theatre and the necessary intraoperative resources. Several studies suggested a staged SLNB procedure prior to mastectomy when IBR is planned^34–36^. While enabling a complete pathological evaluation of the SLN status before breast surgery, this strategy comes with the drawback of a two-step surgical procedure, including increased risk for infections, delay in definitive surgery, and increased arm morbidity and hospital costs^37, 38^. Moreover, the prevalence of lymphatic metastasis at the time of diagnosis is declining^39^, and the number of patients benefitting from an altered treatment plan due to surgically verified macro-SLNM and the need for PMRT is limited^40^. Therefore, a complementary, non-invasive prediction tool would be beneficial to preoperatively identify patients for whom IBR is associated with a high risk of postoperative complications due to nodal macrometastasis and PMRT. Ultimately, evaluating the risk factors impairing the outcome of IBR, including the need for PMRT, is essential to support breast cancer patients and healthcare providers to make informed decisions on breast reconstructive options.

In this study, we present two nomograms for preoperative prediction of SLN status indicating the need for PMRT based on routinely accessible clinicopathological characteristics. The nomograms were well-calibrated and displayed AUC values of 0.708 and 0.740 in the temporal validation cohort. For the prediction of axillary nodal status in cN0 breast cancer, the Memorial Sloan Kettering Cancer Center (MSKCC) nomogram^18^ is one of the most frequently used, with an AUC of 0.754. However, this nomogram includes lymphovascular invasion, which is difficult to obtain in a preoperative setting^41^. Similar to most other nomograms for the prediction of axillary nodal status, it does not consider the size of the metastatic deposit. Previous trials have failed to prove any benefit of PMRT for patients with only micrometastatic deposit (≤2.0 mm)^42, 43^ and micrometastatic disease is not included in the current guidelines concerning recommendations for PMRT^12–14^. Therefore, the presented Nomogram I was developed to predict ≥1 macro-SLNMs specifically. The endpoint of the second nomogram was chosen to adhere to the protocols of ongoing randomized clinical trials questioning the benefit of locoregional radiotherapy for patients with low nodal metastatic burden and verified 1-2 macro-SLNMs. In the POSNOC trial^16^, women with T1-T2 tumors and 1-2 macro-SLNMs are randomized to no further axillary treatment vs. regional radiotherapy/axillary dissection. Similarly, the MA.39^17^ and the T-REX^15^ trials investigate the need for regional lymph node radiotherapy in patients with 1-2 macro-SLNMs; however, these trials are limited to patients with ER-positive, HER2-negative breast cancer. Since radiotherapy is associated with an increased risk of arm lymphedema^44^, postmastectomy pain syndrome^45^, and cardiopulmonary disease^46, 47^, a de-escalation of locoregional radiotherapy would not only improve the outcomes of IBR but also reduce morbidity and non-breast cancer mortality for these patients. Clinical guidelines concerning recommendations for PMRT will need to be revised if the omission of locoregional radiotherapy proves to be non-inferior in these patients, and the importance of identifying metastasis limited to 1-2 macro-SLNMs will be diminished. To our knowledge, the presented Nomogram II is the first nomogram to predict >2 macro-SLNMs in patients with breast cancer for whom PMRT will remain recommended.

Adaptive LASSO regression was used to develop the nomograms. Although the LASSO algorithm has proved to be advantageous in the development of prediction models^48, 49^, the adaptive LASSO regression-based nomograms presented in this study did not outperform the prediction models based on backward stepwise regression with bootstrap uniform shrinkage presented in Supplementary material. Moreover, the different algorithms showed only small differences in variable selection. This observation is most likely due to the restricted number of candidate variables recognized as key predictors, and thus, the advantages offered by the adaptive LASSO algorithm were limited.

This study has some limitations. Besides its retrospective nature, all data were obtained from a quality register with incomplete data for some clinicopathological characteristics. However, NKBC is a national, population-based register with high completeness when cross-linked to the Swedish Cancer Register^21^, and good concordance when validated by re-extraction from medical records^50^. Moreover, although many patients were removed from the modeling due to missing data on one or more of the candidate variables, the study still included a large number of observations (*n* = 18 185). Furthermore, this study is limited by the imbalanced distribution of macro-SLNM and >2 macro-SLNMs in the overall study cohort. Nevertheless, this is an important observation, highlighting the small number of patients benefitting from an altered treatment planning due to pathologically verified macro-SLNM and the need for PMRT. A strength of this study is the temporal validation of the nomograms regarding both discriminatory ability and model calibration. Although only variables that could be obtained in a preoperative setting were included as predictors for nodal disease, the included tumor characteristics were based on the definitive pathology result. Core needle biopsy has been shown to accurately evaluate the histological type and grade, hormone receptor status, and HER2 receptor status^41^; however, the differences between pre-and postoperative values are to be expected. Additionally, mammography may over-or underestimate the pathological tumor size^51^ and the presence of multifocality^52^. The results should, therefore, be interpreted with caution. To enable the clinical application of these nomograms, a prospective validation study is necessary. Moreover, it must be noted that the nomogram does not consider other risk factors for postoperative complications of breast reconstructive surgery, such as obesity, smoking, and diabetes.

In conclusion, this study presents nomograms based on routine clinical patient and tumor characteristics for preoperative risk stratification of SLN status indicating the need for PMRT, addressing both current guidelines on the use of PMRT and ongoing clinical trials aiming to de-escalate irradiation. The nomograms could be valuable in preoperatively evaluating the risks and benefits of IBR regarding the need for PMRT, thus supporting individualized decision-making on breast reconstructive surgery.

## Data Availability

The data sets generated and analyzed in the present study are available from the corresponding author upon reasonable request.

## REFERENCES

1. Dauplat J, Kwiatkowski F, Rouanet P, et al. Quality of life after mastectomy with or without immediate breast reconstruction. Br J Surg. 2017;104(9):1197–206.

2. Archangelo SCV, Sabino Neto M, Veiga DF, Garcia EB, Ferreira LM. Sexuality, depression and body image after breast reconstruction. Clinics (Sao Paulo). 2019;74:e883.

3. Padubidri AN, Yetman R, Browne E, et al. Complications of postmastectomy breast reconstructions in smokers, ex-smokers, and nonsmokers. Plast Reconstr Surg. 2001;107(2):342–9; discussion 50-1.

4. Fischer JP, Nelson JA, Kovach SJ, Serletti JM, Wu LC, Kanchwala S. Impact of obesity on outcomes in breast reconstruction: analysis of 15,937 patients from the ACS-NSQIP datasets. J Am Coll Surg. 2013;217(4):656–64.

5. Mortada H, Alwadai A, Bamakhrama B, et al. The Impact of Diabetes Mellitus on Breast Reconstruction Outcomes and Complications: A Systematic Literature Review and Meta-analysis. Aesthetic Plast Surg. 2023.

6. Barry M, Kell MR. Radiotherapy and breast reconstruction: a meta-analysis. Breast Cancer Res Treat. 2011;127(1):15–22.

7. de Boniface J, Coudé Adam H, Frisell A, et al. Long-term outcomes of implant-based immediate breast reconstruction with and without radiotherapy: a population-based study. Br J Surg. 2022;109(11):1107–15.

8. Jagsi R, Momoh AO, Qi J, et al. Impact of Radiotherapy on Complications and Patient-Reported Outcomes After Breast Reconstruction. J Natl Cancer Inst. 2018;110(2):157–65.

9. Shumway DA, Momoh AO, Sabel MS, Jagsi R. Integration of Breast Reconstruction and Postmastectomy Radiotherapy. J Clin Oncol. 2020;38(20):2329–40.

10. Heller DR, Zhuo H, Zhang Y, et al. Surgical Outcomes of Mastectomy with Immediate Autologous Reconstruction Followed by Radiation. Ann Surg Oncol. 2021;28(4):2169–79.

11. Liew B, Southall C, Kanapathy M, Nikkhah D. Does post-mastectomy radiation therapy worsen outcomes in immediate autologous breast flap reconstruction? A systematic review and meta-analysis. J Plast Reconstr Aesthet Surg. 2021;74(12):3260–80.

12. Cardoso F, Kyriakides S, Ohno S, et al. Early breast cancer: ESMO Clinical Practice Guidelines for diagnosis, treatment and follow-up†. Ann Oncol. 2019;30(8):1194–220.

13. Gradishar WJ, Moran MS, Abraham J, et al. Breast Cancer, Version 3.2022, NCCN Clinical Practice Guidelines in Oncology. J Natl Compr Canc Netw. 2022;20(6):691–722.

14. Recht A, Comen EA, Fine RE, et al. Postmastectomy Radiotherapy: An American Society of Clinical Oncology, American Society for Radiation Oncology, and Society of Surgical Oncology Focused Guideline Update. Pract Radiat Oncol. 2016;6(6):e219–e34.

15. Alkner S, de Boniface J, Lundstedt D, et al. Protocol for the T-REX-trial: tailored regional external beam radiotherapy in clinically node-negative breast cancer patients with 1-2 sentinel node macrometastases - an open, multicentre, randomised non-inferiority phase 3 trial. BMJ Open. 2023;13(9):e075543.

16. Goyal A, Mann GB, Fallowfield L, et al. POSNOC-POsitive Sentinel NOde: adjuvant therapy alone versus adjuvant therapy plus Clearance or axillary radiotherapy: a randomised controlled trial of axillary treatment in women with early-stage breast cancer who have metastases in one or two sentinel nodes. BMJ Open. 2021;11(12):e054365.

17. Parulekar WR, Berrang T, Kong I, et al. Cctg MA.39 tailor RT: A randomized trial of regional radiotherapy in biomarker low-risk node-positive breast cancer (NCT03488693). Journal of Clinical Oncology. 2019;37(15_suppl):TPS602-TPS.

18. Bevilacqua JL, Kattan MW, Fey JV, Cody HS, 3rd, Borgen PI, Van Zee KJ. Doctor, what are my chances of having a positive sentinel node? A validated nomogram for risk estimation. J Clin Oncol. 2007;25(24):3670–9.

19. Meretoja TJ, Heikkilä PS, Mansfield AS, et al. A predictive tool to estimate the risk of axillary metastases in breast cancer patients with negative axillary ultrasound. Ann Surg Oncol. 2014;21(7):2229–36.

20. Geng SK, Fu SM, Zhang HW, Fu YP. Predictive nomogram based on serum tumor markers and clinicopathological features for stratifying lymph node metastasis in breast cancer. BMC Cancer. 2022;22(1):1328.

21. Swedish National Quality Registry for Breast Cancer 2021 [cited 19 Jan 2023]. Available from: https://cancercentrum.se/samverkan/cancerdiagnoser/brost/kvalitetsregister/.

22. Edge S. AJCC Cancer Staging Manual, Eight Edition. New York: Springer; 2017.

23. von Elm E, Altman DG, Egger M, Pocock SJ, Gøtzsche PC, Vandenbroucke JP. The Strengthening the Reporting of Observational Studies in Epidemiology (STROBE) Statement: guidelines for reporting observational studies. Int J Surg. 2014;12(12):1495–9.

24. Collins GS, Reitsma JB, Altman DG, Moons KG. Transparent Reporting of a multivariable prediction model for Individual Prognosis Or Diagnosis (TRIPOD): the TRIPOD Statement. Br J Surg. 2015;102(3):148–58.

25. Viale G, Zurrida S, Maiorano E, et al. Predicting the status of axillary sentinel lymph nodes in 4351 patients with invasive breast carcinoma treated in a single institution. Cancer. 2005;103(3):492–500.

26. Farley C, Bassett R, Meric-Bernstam F, et al. To Dissect or Not to Dissect: Can We Predict the Presence of Four or More Axillary Lymph Node Metastases in Postmenopausal Women with Clinically Node-Negative Breast Cancer? Ann Surg Oncol. 2023;30(13):8327–34.

27. Wildiers H, Van Calster B, van de Poll-Franse LV, et al. Relationship between age and axillary lymph node involvement in women with breast cancer. J Clin Oncol. 2009;27(18):2931–7.

28. Hartman J, Ehinger A, Kovács A, et al. [Kvalitetsbilaga för Bröstpatologi (KVAST)] 2022 [cited 5 Apr 2023]. Available from: https://kunskapsbanken.cancercentrum.se/diagnoser/brostcancer/vardprogram/kvalitetsdokument-for--patologi/#chapter--Forfattare-KVAST-gruppen-for-brostpatologi.

29. Shortreed SM, Ertefaie A. Outcome-adaptive lasso: Variable selection for causal inference. Biometrics. 2017;73(4):1111–22.

30. Tibshirani R. Regression Shrinkage and Selection via the Lasso. Journal of the Royal Statistical Society Series B (Methodological). 1996;58(1):267–88.

31. Tibshirani R. The lasso method for variable selection in the Cox model. Stat Med. 1997;16(4):385–95.

32. Fernandez-Felix BM, García-Esquinas E, Muriel A, Royuela A, Zamora J. Bootstrap internal validation command for predictive logistic regression models. The Stata Journal. 2021;21(2):498–509.

33. Wood BC, Levine EA, Marks MW, David LR. Outcomes of immediate breast reconstruction in patients undergoing single-stage sentinel lymph node biopsy and mastectomy. Ann Plast Surg. 2011;66(5):564–7.

34. Teven C, Agarwal S, Jaskowiak N, et al. Pre-mastectomy sentinel lymph node biopsy: a strategy to enhance outcomes in immediate breast reconstruction. Breast J. 2013;19(5):496–503.

35. McGuire K, Rosenberg AL, Showalter S, Brill KL, Copit S. Timing of sentinel lymph node biopsy and reconstruction for patients undergoing mastectomy. Ann Plast Surg. 2007;59(4):359–63.

36. Klauber-Demore N, Calvo BF, Hultman CS, et al. Staged sentinel lymph node biopsy before mastectomy facilitates surgical planning for breast cancer patients. Am J Surg. 2005;190(4):595–7.

37. Husen M, Paaschburg B, Flyger HL. Two-step axillary operation increases risk of arm morbidity in breast cancer patients. Breast. 2006;15(5):620–8.

38. Rönkä R, Smitten K, Sintonen H, et al. The impact of sentinel node biopsy and axillary staging strategy on hospital costs. Ann Oncol. 2004;15(1):88–94.

39. Tabar L, Chen TH, Yen AM, et al. Effect of Mammography Screening on Mortality by Histological Grade. Cancer Epidemiol Biomarkers Prev. 2018;27(2):154–7.

40. Svensson M, Dihge L. The Role of Surgical Axillary Staging Prior to Immediate Breast Reconstruction in the Era of De-Escalation of Axillary Management in Early Breast Cancer. J Pers Med. 2022;12(8).

41. Rakha EA, Ellis IO. An overview of assessment of prognostic and predictive factors in breast cancer needle core biopsy specimens. J Clin Pathol. 2007;60(12):1300–6.

42. Luo H, Yang OO, He JL, Lan T. Impact of Post-Mastectomy Radiation Therapy for Sentinel Lymph Node Micrometastases in Early-Stage Breast Cancer Patients. Med Sci Monit. 2022;28:e933275.

43. Mamtani A, Patil S, Stempel M, Morrow M. Axillary Micrometastases and Isolated Tumor Cells Are Not an Indication for Post-mastectomy Radiotherapy in Stage 1 and 2 Breast Cancer. Ann Surg Oncol. 2017;24(8):2182–8.

44. Konishi T, Tanabe M, Michihata N, et al. Risk factors for arm lymphedema following breast cancer surgery: a Japanese nationwide database study of 84,022 patients. Breast Cancer. 2023;30(1):36–45.

45. Ren Y, Kong X, Yang Q, et al. Incidence, risk factors, prevention and treatment of postmastectomy pain syndrome in breast cancer: A multicenter study. Int J Surg. 2022;106:106937.

46. Taylor C, Correa C, Duane FK, et al. Estimating the Risks of Breast Cancer Radiotherapy: Evidence From Modern Radiation Doses to the Lungs and Heart and From Previous Randomized Trials. J Clin Oncol. 2017;35(15):1641–9.

47. Darby SC, Ewertz M, McGale P, et al. Risk of ischemic heart disease in women after radiotherapy for breast cancer. N Engl J Med. 2013;368(11):987–98.

48. Su YR, Buist DS, Lee JM, et al. Performance of statistical and machine learning risk prediction models for surveillance benefits and failures in breast cancer survivors. Cancer Epidemiol Biomarkers Prev. 2023.

49. Huang X, Xu X, Xu A, et al. Exploring the most appropriate lymph node staging system for node-positive breast cancer patients and constructing corresponding survival nomograms. J Cancer Res Clin Oncol. 2023.

50. Löfgren L, Eloranta S, Krawiec K, Asterkvist A, Lönnqvist C, Sandelin K. Validation of data quality in the Swedish National Register for Breast Cancer. BMC Public Health. 2019;19(1):495.

51. Gruber IV, Rueckert M, Kagan KO, et al. Measurement of tumour size with mammography, sonography and magnetic resonance imaging as compared to histological tumour size in primary breast cancer. BMC Cancer. 2013;13:328.

52. Steinhof-Radwańska K, Lorek A, Holecki M, et al. Multifocality and Multicentrality in Breast Cancer: Comparison of the Efficiency of Mammography, Contrast-Enhanced Spectral Mammography, and Magnetic Resonance Imaging in a Group of Patients with Primarily Operable Breast Cancer. Curr Oncol. 2021;28(5):4016–30.

