## Supplementary for "Development and Validation of Prediction Models for Sentinel Lymph Node Status Indicating Postmastectomy Radiotherapy in Breast Cancer: a Population-Based Study of 18 185 Women"

### Supplementary Material

#### Supplemental Methods

All candidate predictors were entered in a backward stepwise logistic regression analysis using $P < .157$ as the threshold for variable selection. Cases with missing values of ≥1 candidate predictor were removed from the analyses (Table S2). Two models were developed for predicting ≥1 sentinel lymph node macrometastases (macro-SLNMs) and >2 macro-SLNMs, respectively, based on the results from the logistic regression analysis. Using bootstrapping with 1000 replicates[1], a uniform shrinkage factor was calculated to shrink each coefficient in the two models, minimizing the risk of overfitting. To evaluate the discriminatory ability for each model, the area under the receiver operating characteristic (ROC) curve (AUC) was calculated. Model calibration was assessed graphically using calibration plots and numerically as calibration slope and intercept.

#### Supplemental Results

In the multivariable logistic regression analysis, variables that were retained in the final model for ≥1 macro-SLNMs were age, tumor size, histological type and grade, ER status, and multifocality (Table S3). Using bootstrapping with 1000 replicates, the coefficients were modified with a uniform shrinkage factor of 0.988. For the prediction of >2 macro-SLNMs, tumor size, histological grade, and multifocality were retained in the final model after backward stepwise regression. A uniform shrinkage factor of 0.952 was applied to these regression coefficients using bootstrapping.

ROC curves and calibration plots illustrating the predictive performance and accuracy of the two prediction models based on backward stepwise regression with bootstrap uniform shrinkage are presented in Fig. S3. The AUC value for the model predicting ≥1 macro-SLNMs was estimated to be 0.719 (95% confidence interval, 0.707–0.732) in the training cohort and 0.703 (0.680–0.727) in the validation cohort. The model was well-calibrated with a calibration slope and intercept of 1.017 and -0.113, respectively, in the validation cohort. For the model predicting the presence of >2 macro-SLNMs, the AUC in the training and validation cohort was estimated to be 0.773 (0.740–0.805) and 0.742 (0.684–0.800), respectively. Similarly, the model was well-calibrated with calibration slope and calibration intercept of 1.015 and 0.162, respectively.

#### Supplementary Tables

##### Table S1. Univariable logistic regression analyses of clinicopathological characteristics predictive of ≥1 and >2 sentinel lymph node macrometastases (macro-SLNMs) (*n* = 13 656)

| **Variables** | **Prediction of ≥1 macro-SLNMs** | | **Prediction of >2 macro-SLNMs** | |
|---|---|---|---|---|
| | **OR (95% CI)** | ***P* value** | **OR (95% CI)** | ***P* value** |
| **Age** (years) | | | | |
| ≤65 | 1 (reference) | | 1 (reference) | |
| 66-75 | 0.709 (0.633 to 0.795) | <.001 | 0.767 (0.553 to 1.064) | .112 |
| >75 | 1.254 (1.090 to 1.444) | .002 | 1.476 (1.017 to 2.142) | .040 |
| **Tumor size** (mm) | 1.070 (1.064 to 1.075) | <.001 | 1.085 (1.073 to 1.098) | <.001 |
| **Histological type** | | | | |
| NST | 1 (reference) | | 1 (reference) | |
| ILC | 1.236 (1.077 to 1.418) | .003 | 2.223 (1.600 to 3.087) | <.001 |
| Others | 0.437 (0.337 to 0.568) | <.001 | 0.795 (0.417 to 1.516) | .486 |
| **Histological grade** | | | | |
| I | 1 (reference) | | 1 (reference) | |
| II | 2.066 (1.779 to 2.398) | <.001 | 3.757 (2.156 to 6.548) | <.001 |
| III | 2.599 (2.212 to 3.053) | <.001 | 4.474 (2.508 to 7.981) | <.001 |
| **Multifocality** (multifocal *vs.* unifocal) | 2.043 (1.818 to 2.296) | <.001 | 1.941 (1.417 to 2.658) | <.001 |
| **ER status** (pos *vs.* neg) | 1.072 (0.887 to 1.295) | .473 | 0.893 (0.541 to 1.475) | .658 |
| **PR status** (pos *vs.* neg) | 1.084 (0.936 to 1.255) | .280 | 0.859 (0.583 to 1.266) | .442 |
| **HER2 status** (pos *vs.* neg) | 1.112 (0.954 to 1.295) | .174 | 1.312 (0.876 to 1.963) | .187 |

Binary coding was used for categorical variables with two levels and two binary so-called dummy variables for categorical variables with three levels.

*OR*, odds ratio; *CI*, confidence interval; *NST*, ductal carcinoma of no special type; *ILC*, invasive lobular carcinoma; *ER*, estrogen receptor; *PR*, progesterone receptor; *HER2*, human epidermal growth factor receptor 2.

**Table S2. Patient and tumor characteristics of those included in the multivariable analyses and those removed due to any missing value of the candidate predictors**

| **Variable** | **Training cohort** ***n*** **= 13 656** | **Included cases** ***n*** **= 12 168** | **Removed cases** ***n*** **= 1488** |
|---|---|---|---|
| **Age** (years), median (range) | 54 (23-95) | 64 (23-95) | 65 (25-94) |
| Missing | 0 | 0 | 0 |
| **Patient age,** categories | | | |
| ≤65 years | 7320 (53) | 6528 (54) | 792 (53) |
| 66-75 years | 4626 (34) | 4142 (34) | 484 (33) |
| >75 years | 1710 (13) | 1498 (12) | 212 (14) |
| Missing | 0 | 0 | 0 |
| **Tumor size** (mm), median (range) | 15 (1-50) | 15 (1-50) | 15 (1-50) |
| Missing | 0 | 0 | 0 |
| **Histological type** | | | |
| NST | 10 511 (79) | 9652 (79) | 859 (78) |
| ILC | 1779 (13) | 1672 (14) | 107 (10) |
| Others | 977 (7) | 844 (7) | 133 (12) |
| Missing | 389 | 0 | 389 |
| **Nottingham histological grade** | | | |
| I | 3081 (23) | 2840 (23) | 241 (18) |
| II | 7118 (53) | 6466 (53) | 652 (49) |
| III | 3298 (24) | 2862 (24) | 436 (33) |
| Missing | 159 | 0 | 159 |
| **Multifocality** | | | |
| Yes | 2167 (16) | 1846 (15) | 321 (22) |
| No | 11 463 (84) | 10 322 (85) | 1141 (78) |
| Missing | 26 | 0 | 26 |
| **ER status** | | | |
| Positive | 12 032 (92) | 11 193 (92) | 839 (94) |
| Negative | 1028 (8) | 975 (8) | 53 (6) |
| Missing | 596 | 0 | 596 |
| **PR status** | | | |
| Positive | 10 988 (86) | 10 416 (86) | 572 (86) |
| Negative | 1844 (14) | 1752 (14) | 92 (14) |
| Missing | 824 | 0 | 824 |
| **HER2 status** | | | |
| Positive | 1497 (11) | 1290 (11) | 207 (16) |
| Negative | 11 939 (89) | 10 878 (89) | 1061 (84) |
| Missing | 220 | 0 | 220 |
| **≥1 macro-SLNMs** | | | |
| Yes | 1852 (14) | 1647 (14) | 205 (14) |
| No | 11 804 (86) | 10 521 (86) | 1283 (86) |
| Missing | 0 | 0 | 0 |
| **>2 macro-SLNMs** | | | |
| Yes | 203 (1) | 175 (1) | 28 (2) |
| No | 13 453 (99) | 11 993 (99) | 1460 (98) |
| Missing | 0 | 0 | 0 |

Values in parentheses are valid percentages of each column if not otherwise explained. The percentage values are rounded, and the total percentage may, therefore, not be 100.

*NST*, no special type; *ILC*, invasive lobular cancer; *ER*, estrogen receptor; *PR*, progesterone receptor; *HER2*, human epidermal growth factor receptor 2; *macro-SLNMs*, sentinel lymph node macrometastases.

**Table S3. Regression coefficients of the variables in the two prediction models based on backward stepwise regression with bootstrap uniform shrinkage (*n* = 12 168)**

| **Variable** | **Prediction of ≥1 macro-SLNMs** | | **Prediction of >2 macro-SLNMs** | |
|---|---|---|---|---|
| | **Shrinkage coefficients (shrinkage factor = 0.988)** | ***P* value** | **Shrinkage coefficients (shrinkage factor = 0.952)** | ***P* value** |
| **Age** (years) | | | | |
| ≤65 | 0 (reference) | | | |
| 66-75 | -0.276 (-0.400 to -0.152) | <.001 | | |
| >75 | -0.131 (-0.290 to 0.029) | .108 | | |
| **Tumor size** (mm) | 0.067 (0.061 to 0.073) | <.001 | 0.072 (0.060 to 0.084) | <.001 |
| **Histological type** | | | | |
| NST | 0 (reference) | | | |
| ILC | -0.266 (-0.425 to -0.107) | .001 | | |
| Others | -0.651 (-0.933 to -0.369) | <.001 | | |
| **Histological grade** | | | | |
| I | 0 (reference) | | 0 (reference) | |
| II | 0.351 (0.187 to 0.514) | <.001 | 0.837 (0.258 to 1.416) | .005 |
| III | 0.461 (0.276 to 0.656) | <.001 | 0.900 (0.295 to 1.505) | .004 |
| **ER status** (pos *vs* neg) | 0.332 (0.119 to 0.0.546) | .002 | | |
| **Multifocality** (multifocal *vs* unifocal) | 0.635 (0.505 to 0.766) | <.001 | 0.505 (0.172 to 0.838) | .003 |
| **Constant** | -3.643 | | -6.508 | |

The constants and coefficients, estimated by the logistic regression algorithm, constitute the function used for predicting the outcome. A positive coefficient indicates that the variable increases the predicted probability and a negative coefficient indicates that the variable decreases the predicted probability. Using bootstrapping, the coefficients have been shrunken to minimize the risk of overfitting. Binary coding was used for categorical variables with two levels and two binary so-called dummy variables for categorical variables with three levels.

*Macro-SLNMs*, sentinel lymph node macrometastases; *NST*, ductal carcinoma of no special type; *ILC*, invasive lobular cancer; *ER*, estrogen receptor.

**Supplementary Figures**

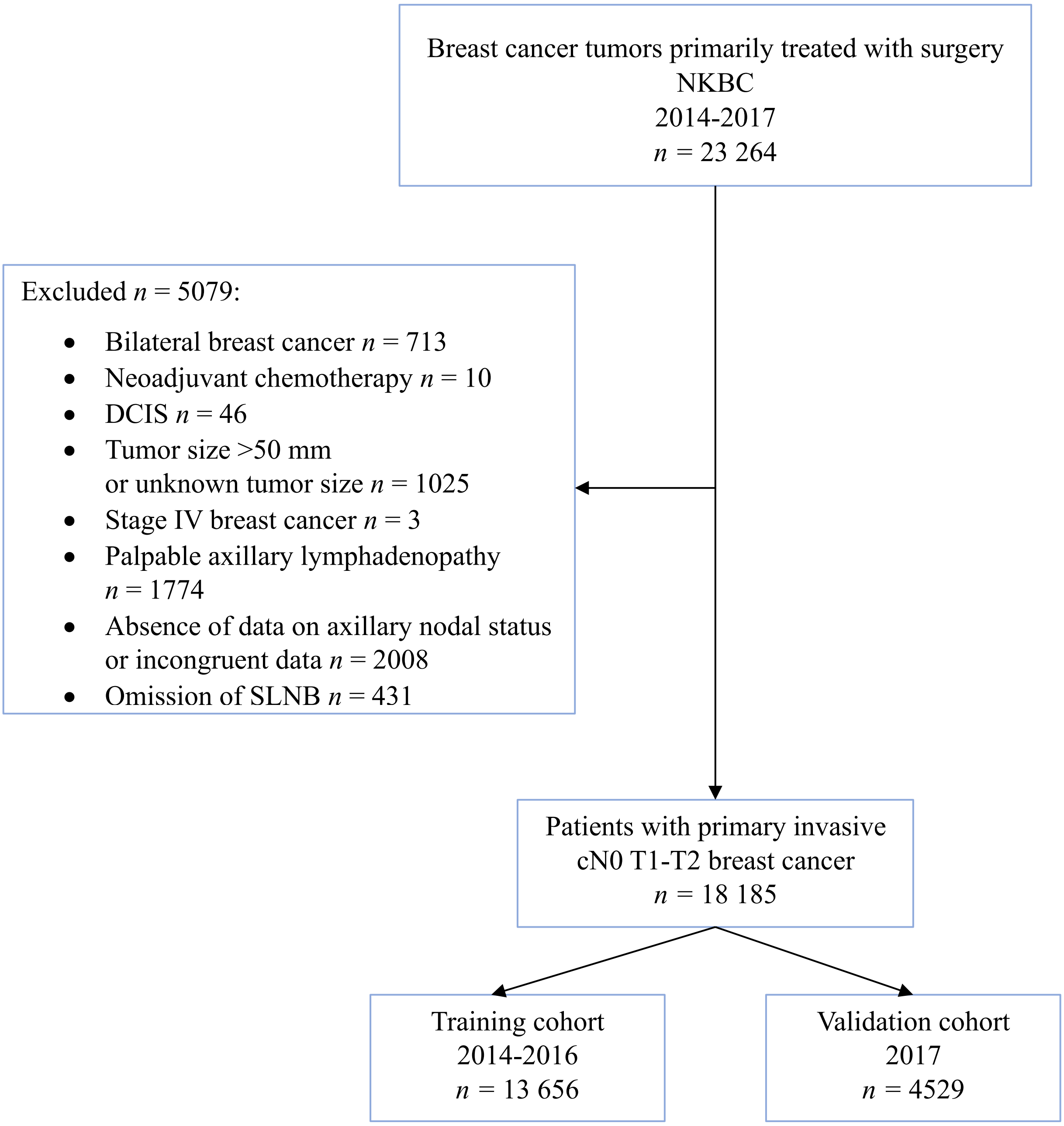


**Figure S1. Flow chart of the study cohort.**

*NKBC*, Swedish National Quality Registry for Breast Cancer; *DCIS*, ductal carcinoma *in situ*; *SLNB*, sentinel lymph node biopsy; *cN0*, clinically node-negative.

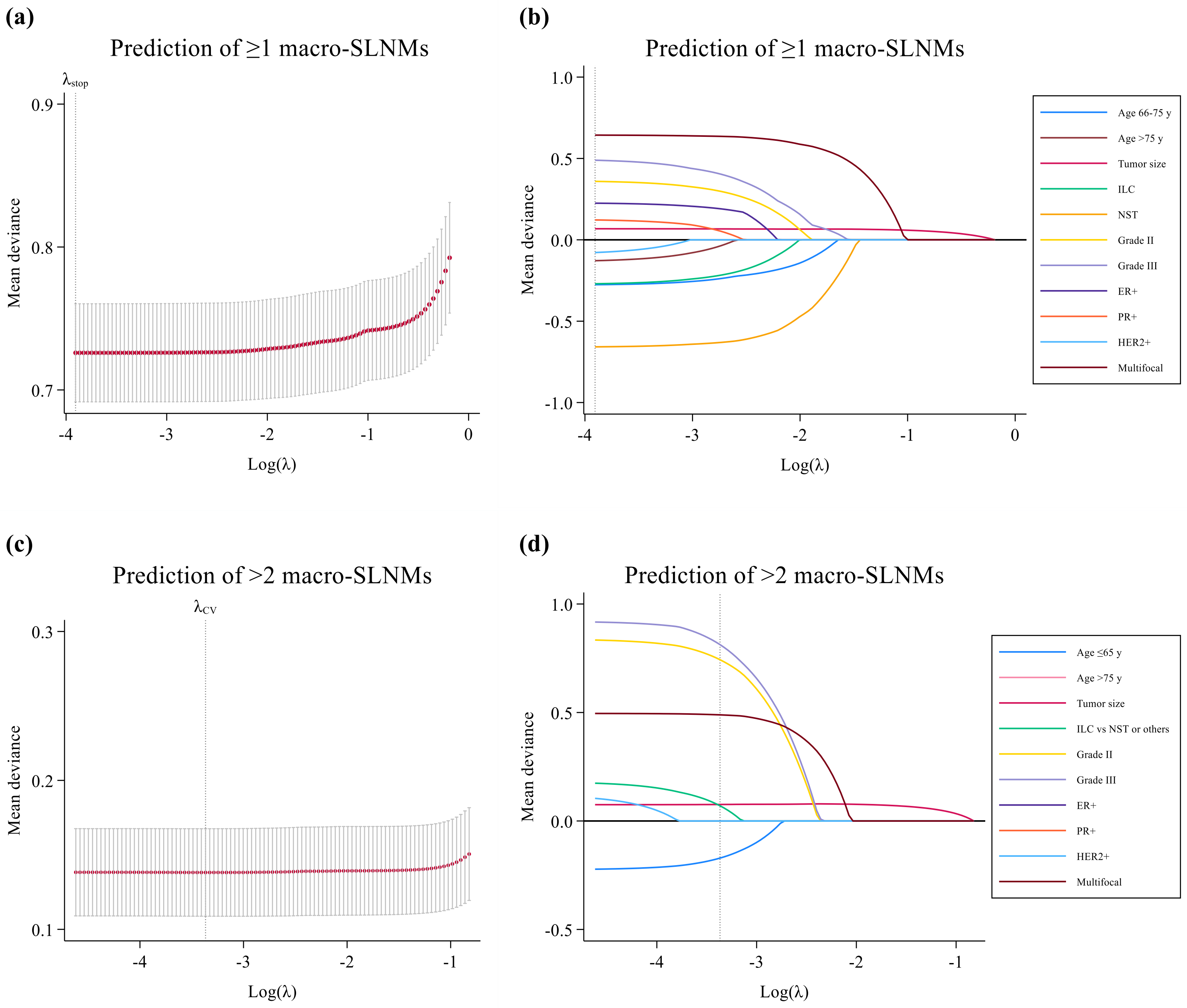


**Figure S2. Adaptive LASSO logistic regression analyses for prediction of ≥1 and >2 sentinel lymph node macrometastases (macro-SLNMs).** The adaptive LASSO regression algorithm forces the absolute values of the regression coefficients of the standardized predictors to be bounded by a penalty factor λ. Here, the optimal value of λ was determined using 10-fold cross-validation (CV) to minimize the mean deviation across the CV models. Variables with a non-zero coefficient for the optimal value of $\lambda$ were selected for the prediction models. (**a**) A λ value of 0.00012, log(λ) = -3.90588 identified (**b**) 11 non-zero coefficients for prediction of ≥1 macro-SLNMs, and (**c**) a λ value of 0.00043, log(λ) = -3.36683 identified (**d**) 6 non-zero coefficients for prediction of >2 macro-SLNMs.

*ILC,* invasive lobular carcinoma; *NST,* no special type; *ER+*, estrogen receptor-positive; *PR+,* progesterone receptor positive; *HER2+,* human epidermal growth factor receptor positive.

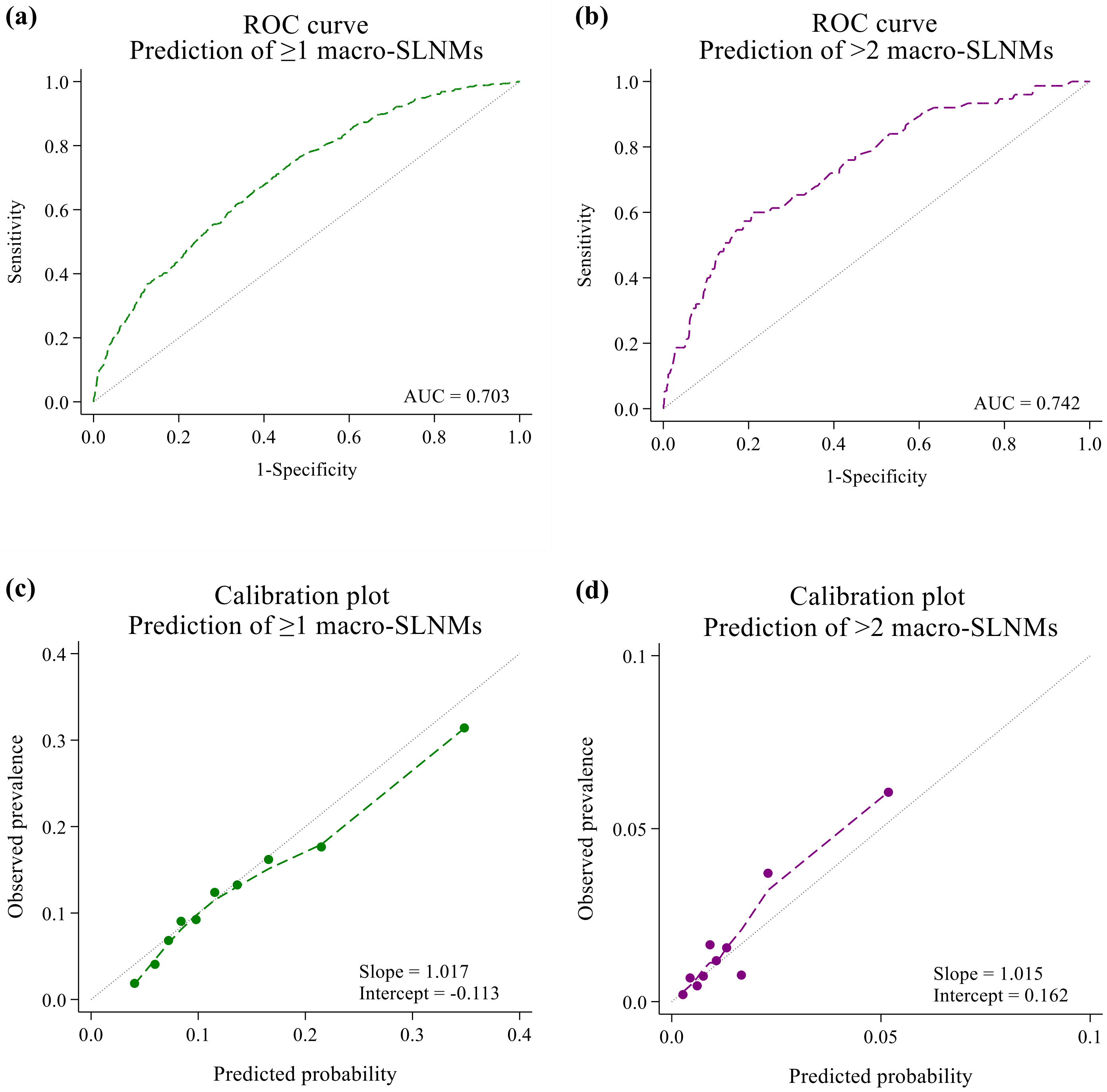


**Figure S3. Performance of the prediction models based on backward stepwise regression with bootstrap uniform shrinkage in the temporal validation cohort.** Receiver operating characteristic (ROC) curves representing the discriminatory ability for (**a**) prediction of ≥1 sentinel lymph node macrometastases (macro-SLNMs) and (**b**) prediction of >2 macro-SLNMs.

The Calibration plots illustrate the agreement between the observed prevalence and the predicted probability of (**c**) ≥1 macro-SLNMs and (**d**) >2 macro-SLNMs, respectively.

*AUC,* area under the curve.

#### SUPPLEMENTARY REFERENCE

1. Fernandez-Felix BM, García-Esquinas E, Muriel A, Royuela A, Zamora J. Bootstrap internal validation command for predictive logistic regression models. The Stata Journal. 2021;21(2):498-509.
